# A Supervised Machine Learning Model to Predict Therapy Response and Mortality at 90 days After Acute Myeloid Leukemia Diagnosis

**DOI:** 10.1101/2023.06.26.23291731

**Authors:** Juan A. Delgado Sanchis, Pedro Pons-Suñer, Noemi Alvarez, Claudia Sargas, Sara Dorado, Jose Vicente Gil Ortí, François Signol, Marta Llop, Laura Arnal, Rafael Llobet, Juan-Carlos Perez-Cortes, Rosa Ayala, Eva Barragán

## Abstract

**Background and Objective:** The main objective in this paper is to validate a machine-learning model trained to predict the 90-day risk of complications for patients with Acute Myeloid Leukemia using variables available at diagnosis. This is a first fundamental step towards the development of a tool that could help physicians in their therapeutic decisions.

**Methods:** 266 patients and 36 variables form the training dataset collected by Hospital 12 de Octubre (Madrid, Spain). The external test cohort provided by Instituto de Investigación Sanitaria La Fe (Valencia, Spain) contains 162 observations. An XGBoost model was trained with one dataset and validated with the other. Additionally, the features were ranked by permutation importance and compared with the ELN 2022 risk classification by genetics at initial diagnosis.

**Results:** The model was evaluated with the training cohort using leave-one-out cross-validation, reaching a ROC-AUC of 0.85. By setting the functioning point that maximises Youden’s index, 3 out of 4 patients with complications and 84 out of 100 in remission are correctly classified. The model was validated with external data collected in a different hospital, achieving 0.7 ROC-AUC. At the best functioning point, almost 6 out of 10 patients with complications and 8 out of 10 patients in remission are correctly classified. Ranking the variables by descending importance, the top four are, in order: age, white-blood-cells count, Gender, and TP53. The list exhibits good coherence with the ELN 2022 risk classification.

**Conclusions:** The model achieves performances that suggest it could be used as a therapeutical decision support tool. Important variables are coherent with ELN 2022 risk classification. Further work is needed to understand the reasons for the drop in test performance. The 90-day model should be supplemented by others that predict the risk of complications at six months or one year.

## 1. Introduction

Acute myeloid leukemia (AML) is a heterogeneous disease characterized by a wide range of molecular alterations leading to malignant transformation of hematopoietic stem cells. (Sargas, C. Cancers 2022). It is the most frequent type of acute leukemia in adults with a 5-year relative survival of 30.5% reported by Surveillance, Epidemiology, and End Results Program (SEER) in USA (SEER12: https://seer.cancer.gov/statfacts/html/amyl.html). The European Cancer Information System (ECIS) reported a 5-year relative survival of 46.9% for men aged 15 to 44 years versus 7.5% for men aged 65-74 years (Source: ECIS - European Cancer Information System From https://ecis.jrc.ec.europa.eu, accessed on 3/JAN/2023).

The goal of treatment is to achieve, whenever possible, complete remission (CR) of leukemia with initial therapy, followed by consolidation and/or maintenance efforts to deepen the remission and maximize response duration. The induction therapy with one or two cycles of anthracyclines and cytarabine remains the backbone for fit patients for intensive chemotherapy. After attainment of CR, patients are consolidated ideally with regimens that include intermediate-dose cytarabine Döhner et al. (2022). Hypomethylating or low doses of cytarabine combination therapy (with venetoclax, ivosidenib, and other investigational agents) are the most frequent schemes used for patients not fit for intensive treatment.

Treatment failure occurs due to initial resistance and failure to achieve CR but more frequently due to the recurrence of leukemia after achieving CR. In addition, treatment toxicity contributes to the mortality of these patients, especially in the early phases of treatment.

Therefore, evaluating the response is very important to adjust the treatment. Then, non-fit intensive treatment patients should be evaluated early during the first cycle, after three cycles, and then repeated every three cycles for patients in remission Döhner et al. (2022) and fit intensive after the first cycle, and every two cycles for patients in remission.

This paper presents a machine learning-based predictive model that estimates the 90-day risk of complications (or equivalently the chances of treatment success) in patients diagnosed with acute myeloid leukemia (AML) based on the biological characteristics of the patient at diagnosis in order to be able to adjust the initial treatment, avoiding probable unnecessary toxicities. The training population was acquired by Hospital 12 de Octubre in Madrid. The test population was acquired by the Hospital La Fe in Valencia and, consequently, is an external cohort collected in other conditions which is used to estimate the degree of generalization achieved by the trained model. It gives an idea about the performance that could be achieved with data of patients from other centers.

Figure 1 describes the question the predictive model responds to. After being AML-diagnosed, during the pre-induction phase, some data of each patient are collected and the model can be consulted, returning the risk of suffering complications during the next 90 days. Complications are understood as resistant disease, recurrence or death. As it can be seen, although the results of this paper correspond to a 90-day prediction, the approach is flexible and allows us to adjust the prediction window to the duration(s) that are useful for therapeutic decision-making (6 months, 1 year, etc.). To anticipate the evolution of patients in the short, medium or long term, it is sufficient to train a new model that would complement the one set at 90 days.

**Figure 1:**
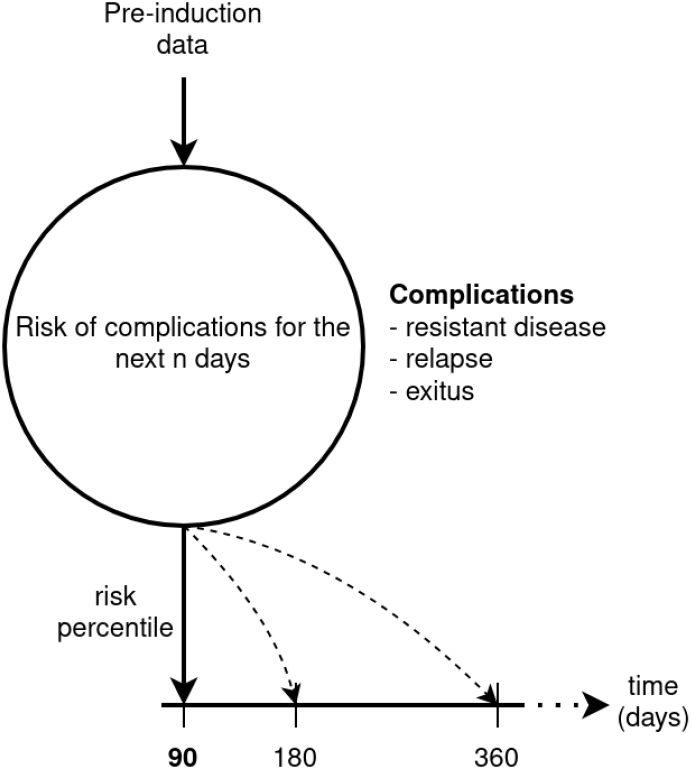
*n*-days risk of complications for AML patients.

Previous studies have demonstrated the usefulness of AI models in predicting the evolution of patients with AML. Gerstung et al. Gerstung et al. (2017) showed that IA models could tailor the AML therapeutic decisions according to specific patient needs. The model they used is based on survival analysis (log-additive Cox hazard modeling) and provides the temporal evolution of a patient starting from the diagnosis providing the probabilities a patient has to be alive with or without remission and with or without recurrence with elapsed time. Siddiqui et al. Siddiqui et al. (2022) compared several ML models (Logistic-regression, Decision-Tree and Random-Forest) in predicting in-hospital mortality after induction with data available at diagnosis. The models achieved ROC-AUC scores ranging from 0.70 to 0.78 and could have helped to detect 51 patients who suffered treatment-related mortality. Another comparison of 9 ML models is presented in Eckardt et al. (2023) where they were trained for two purposes: predict complete remission and 2-year overall survival. In an external test cohort, models achieved ROC-AUC ranging in [0.71-0.80] and [0.65-0.75], respectively.

Section 2 describes the dataset and how it was acquired and preprocessed. Section 3 presents the performance exhibited by the predictive model. Section 4 is a discussion about the main findings of this paper. Finally, conclusions and future works are detailed in section 5 and section 6, respectively.

## 2. Material and methods

### 2.1. Training dataset

The *Comité de Ética de la Investigación Clínica con Medicamentos* (CEIm) of the *Hospital 12 de Octubre* (Madrid, Spain) approved this work with registration number 19/434. The dataset was collected by Hospital 12 de Octubre and comprises 500 patients and 151 variables. After filtering out patients and variables with insufficient quality, the training dataset is formed with 266 individuals and 36 variables. The quality criteria are described in sections 2.3 and 2.4.

The variables are grouped into the following categories: demographics, preinduction analytics, cytogenetics, and VAF genetics.

Cytogenetic variables are binary, where 1 means the chromosome is altered while 0 means it is normal. Variant Allele Frequency genetics variables are continuous between 0 and 1. They correspond to the percentage of mutated cells observed in a sample.

### 2.2. Validation dataset

The dataset was acquired by *Instituto de Investigación Sanitaria La Fe* (IIS-LaFe, Valencia, Spain). All adult patients (*>* 18 years) with newly diagnosed or relapsed/refractory AML (excluding acute promyelocytic leukemia) according to the World Health Organization criteria (2016 and 2022), regardless of treatment received, were eligible for the study. The Institutional Ethics Committee for Clinical Research of IISLaFe approved this study with registration number 2019/0117. Written informed consent in accordance with the recommendations of the Declaration of Human Rights, the Conference of Helsinki, and institutional regulations were obtained from all patients.

It contains 221 patients with 23 variables among the 36 variables which were used during the training process. The remaining 13 unavailable features are automatically imputed using the same imputation methods applied during the training data pre-processing. Some patients are excluded according to the quality criteria defined in section 2.4, which results in an external test cohort of 162 patients and 36 variables (of which 13 are imputed).

### 2.3. Data curation

A data curation step was carried out to define the variables that can enter the predictive model. For this purpose, several criteria are taken into account:

- Bias criterion: variables exhibiting a bias must be discarded. For instance, a variable with a strong correlation with the target must be revised and, if this correlation is due to an abnormality during data acquisition, the feature must be removed.
- Quality criterion: variables with missing values that cannot be imputed correctly, variables that are quasi-constants (for example, a binary variable where only a small number of patients have values distinct from the majority (mode)).
- Expert criterion: if it is already known, or there is a suspicion about certain variables being influential in the evolution of leukemia, they should be included while their quality is sufficient.

The results of the quality check are contrasted with health professionals. Their expertise allows to confirm which variables are included in the model and which are ultimately discarded. The 36 variables passing the quality criteria are detailed in the Results chapter section 3.1.

#### 2.3.1. Quasi-constant variables

The quasi-constant variables are eliminated, concretely, those where the most frequent value (mode) is present in more than 95% of the patients. In this case, a variable where less than 13 patients have a different value from the mode is discarded. This preprocessing step is referred to as quasi-constant filtering.

These variables cannot be taken into account due to the lack of observations (reduced N), which makes it impossible to determine whether their possible relation with the target is objective or due to chance. In other words, to disambiguate these quasi-constant variables, increasing the dataset’s N is necessary.

#### 2.3.2. Missing values

Several variables with many missing data were discarded from the study due to the uncertainty that may exist in imputing them. First, the optimal imputation method for each feature is automatically chosen from a list of univariate (mean, mode and zero imputing) and multivariate imputers (Iterative Random Forest and Bayesian Ridge Regression), according to the best imputation performance on cross-validation of the training set. Then, taking into account both the fraction of missing entries and the quality of imputation for each variable, those with more than 40% missing values that cannot be satisfactorily imputed are filtered out.

#### 2.3.3. Correlated variables

Correlated variables generally do not improve models and could mask valuable interactions between features when using a tree-based algorithm, as well as perturbing feature importance estimations Genuer et al. (2010). We find that the maximum absolute Pearson correlation coefficient between any pair of features is below 0.6. However, as some sets of features in our data tend to be coincidentally missing in the same patients, the imputation process can introduce correlation between those variables. For this reason, a post-imputation filter of correlated variables is performed with a Pearson coefficient greater than 80%. When two variables have a correlation above 0.8, one is excluded from the study. An exception is made for variables whose influence on AML has already been established. This variables white-list is extracted from the set of genetic abnormalities recognised in the 2022 European LeukemiaNet Döhner et al. (2022). Finally, a last filter ignoring this white list is applied to discard very correlated variables with a threshold of 95% correlation.

### 2.4. 90-day condition labelling

The follow-up of each patient allows us to know their condition (in remission or with complication) 90 days after being diagnosed with Acute Myeloid Leukemia. Therefore, the solution adopted in this paper is based on supervised learning.

The predictive model is built (trained) using both the variables associated with a patient and their condition after 90 days. Its 90-day condition, also called *target*, corresponds to the value that the model should predict. Patients with resistant disease, recurrence or exitus are merged into one single category: *with complication*. These patients are assigned a target value of 1 (positives). Patients in remission belong to *in remission* category and are assigned a target value of 0 (negatives). The prediction task is a binary classification.

To determine the 90-day patient status, the elapsed times between several dates are used: last follow-up, death, recurrence, remission, and diagnosis.

Figure 2 illustrates the rules to establish the 90-day patient status. Patients with complications during the first 90 days are those (1) who decease or (2) with a recurrence (complete remission then recurrence) or (3) with a resistant disease (complete remission later than 90 days). Patients in complete remission and a last follow-up posterior to 90 days are considered as remitted. Some patients with complete remission were discarded due to a last follow-up before the first 90 days. In that situation, they cannot be guaranteed to remain in remission at the 90-day mark.

**Figure 2:**
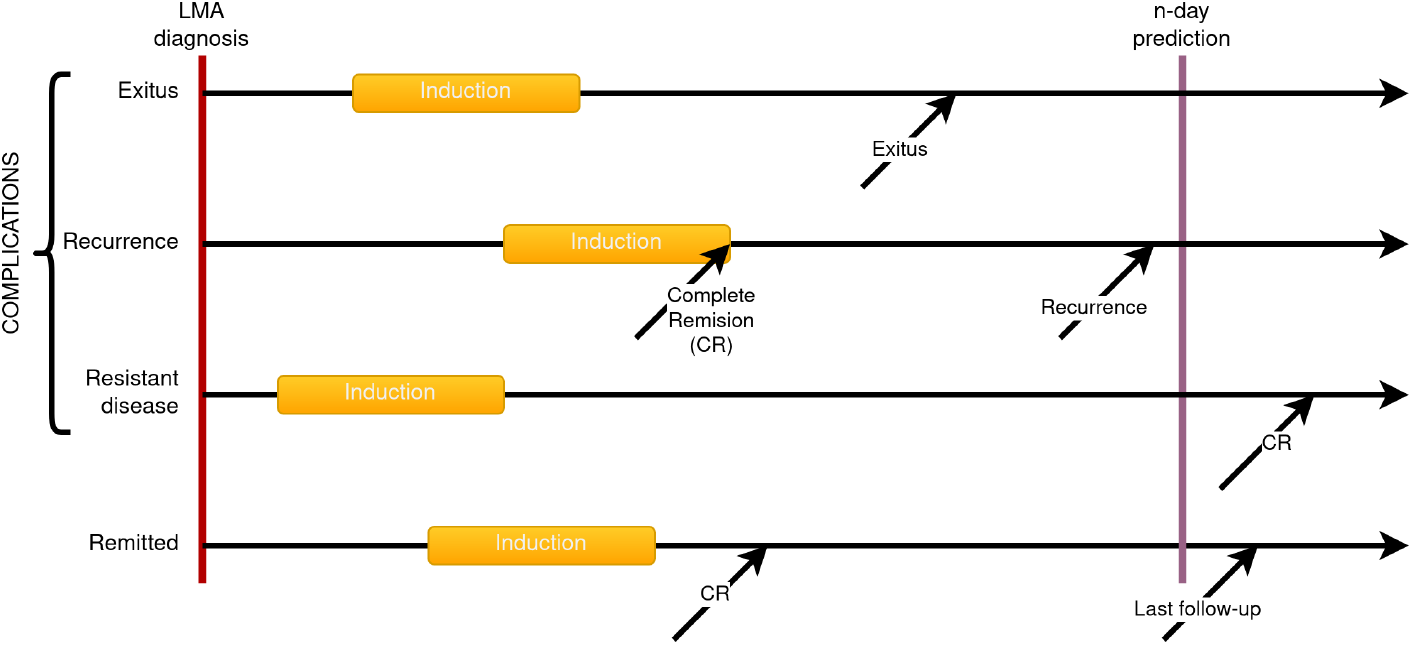
n-day state computing rules. In this study, the prediction is made at 90 days.

Patients whose sequence of dates is inconsistent (missing date, reversed order) are excluded from the study.

Table 1 presents the number of patients for each state in the training and test cohorts. At 90 days after AML diagnosis, the training dataset contains 102 patients with exitus, 32 with recurrence, and 19 with resistant disease forming a population of 153 patients with complications (positives with target 1). 113 patients have entered and remain in complete remission (negatives with target 0). The test dataset comprises 44 patients with exitus, 29 with recurrence, 13 with resistant disease, and 76 in complete remission. In both datasets, classes are balanced as the number of patients with complications (positive) is comparable to those in remission (negative).

**Table 1:** Number of patients in each state for training and test dataset. The class balance %complications/%remitted is provided in the last row.

|  | training | test |
| --- | --- | --- |
| remission | 113 | 76 |
| resistant disease | 19 | 13 |
| recurrence | 32 | 29 |
| exitus | 102 | 44 |
| total | 266 | 162 |
| %positive/%negative | 58/42 | 53/47 |

### 2.5. Predictive model

The predictive model is an Extreme Gradient Boosted Trees (XGBoost) whose use is spreading for clinical decision support in many different diseases as, for example, covid-19 Karthikeyan et al. (2021) Montomoli et al. (2021), coronary diseases Budholiya et al. (2022) or osteosarcoma Jiang et al. (2021).

The model consists of training a set of decision trees in series minimising the error on the prediction of complications, each tree trying to improve where the previous failed. Those interested in further understanding of this model can refer to Hastie et al. (2009) Chen & Guestrin (2016).

In this study, an XGBoost classifier provides the probability that a patient belongs to the “with complications” class or “in remission” class.

### 2.6. Evaluation method and metrics

#### 2.6.1. Cross-validation with a training dataset

Cross-validation by *leave-one-out* is used as the validation method for our model configuration. For each model, a single patient serves as the test and the rest as the training set. The probability of complication for each patient is computed. The ROC curve is drawn by varying a threshold on the probability of having a complication and computing the sensibility and sensitivity at each threshold step.

The ROC curve is enriched with the Youden operating point that globally minimises the errors made by the model. Again, it is important to highlight that it is an example and the final user may choose a different operating point according to a cost (error) matrix or any specific desired behaviour for the model.

#### 2.6.2. Validation with an external dataset

The LAFE external dataset is used to test the model predictions with data other than the one used for training and adjusting the model configuration and where the capture conditions may differ. Each patient’s data from this validation dataset is sent for prediction to the model trained with the H12O dataset, returning their probability of having complications. Then, a ROC curve is obtained.

#### 2.6.3. ROC curve

The classical *Receiver Operating Characteristic* ROC curve shows the true positive rate in function with the false positive rate. The ROC is computed by varying the decision threshold of the model from one extreme, where the model declares that all patients will suffer a complication (maximum sensitivity), to another scenario in which all patients will be in complete remission (maximum specificity). The metrics mentioned above are calculated for each step on the decision threshold.

In this paper, the ROC curve is represented as a sensitivity-sensibility curve. It contains the same information as a classical ROC curve, but it is represented in a way that could be more natural for medical staff to interpret, with indicators they are accustomed to using.

#### 2.6.4. Functioning point

All along the ROC curve, a functioning (operating) point can be set by the final user of the predictive model. By setting this point, the user configures the model in a trade-off between specificity and sensibility.

As an example, in this paper, the operating point is the one that maximizes the Youden’s index. Equation 1 shows the calculation of *Youden’s index*. It is the point that maximises the sum of sensibility and specificity (or minimises the sum of errors) of the model.

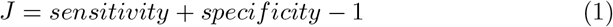

## 3. Results

### 3.1. Final variables set

The data curation process resulted in 36 usable variables presented in Table 2. There are variables of the four categories: demographics, pre-induction analytics, cytogenetics, and genetic abnormalities.

**Table 2:**
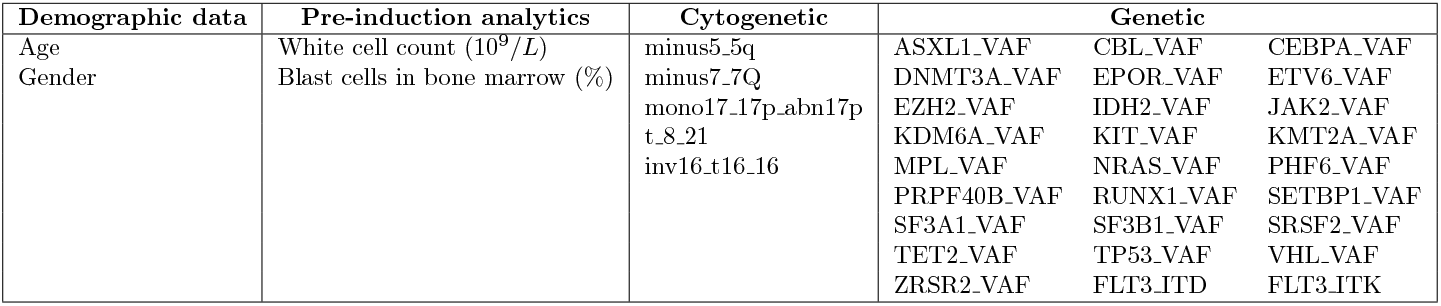
List of the 36 variables that feed the model

### 3.2. Features importance

We can estimate each feature’s importance *for the trained model via* permutation importance (first introduced in Breiman (2001)). To validate feature importances with data from H12O and ensure set-independent results, the permutation-importance algorithm has been performed over multiple validation sets inside a cross-validation with this dataset. Figure 3 presents the top 10 variables sorted by decreasing order of importance.

**Figure 3:**
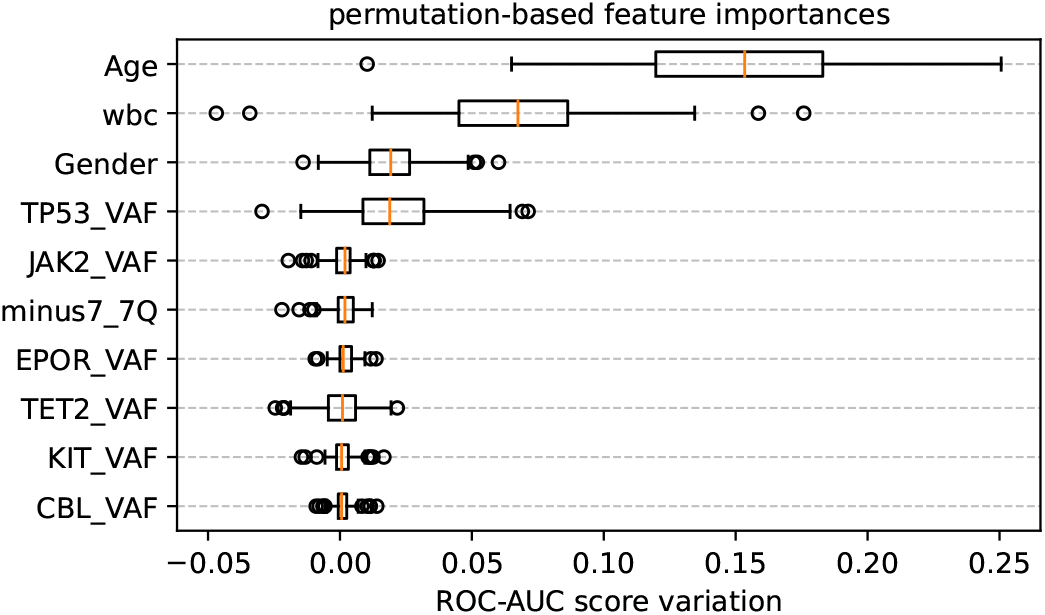
Permutation importance of the ten most important features in the XGB model.

### 3.3. Model evaluation with H12O dataset

Figure 4 shows the sensitivity-specificity curve of the model using a *leaveone-out* cross-validation on the H12O dataset with XGBoost. The area under the ROC curve (AUC) is 0.85. For reference, a model that makes random predictions would get an AUC of around 0.5 with a curve close to the dashed diagonal, while a perfect model would get an AUC of 1 with a perfectly flat curve at 100% sensitivity.

**Figure 4:**
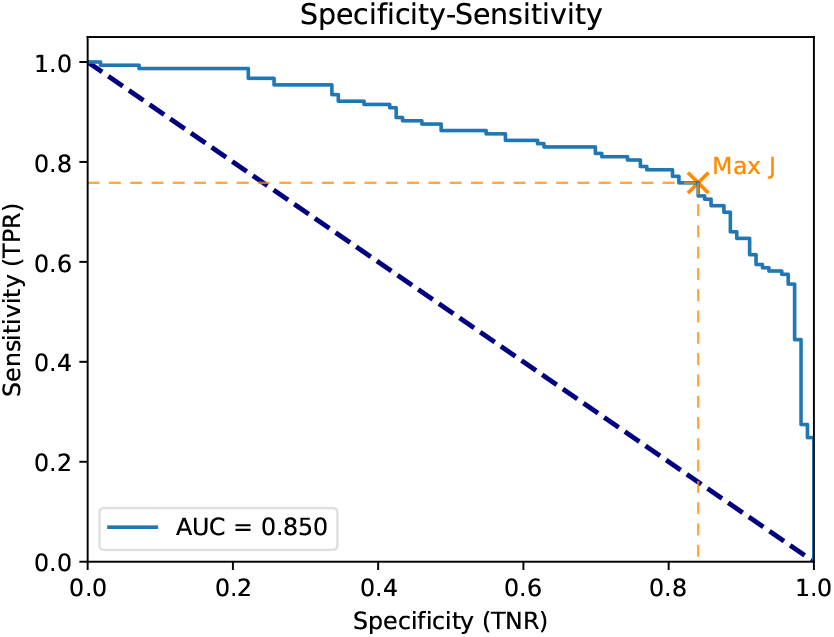
Sensitivity-specificity curve for the training dataset evaluated with leave-one-out cross validation. The operating point is superimposed for maximum Youden’s index (J) at 0.57 a-posteriori probability, percentile 50%.

Table 3 shows the confusion matrix obtained at the operating point that maximizes the Youden’s index (J) in the ROC, displaying basic performance measures such as sensitivity, specificity, positive predictive value (PPV) and negative predictive value (NPV).

**Table 3:**
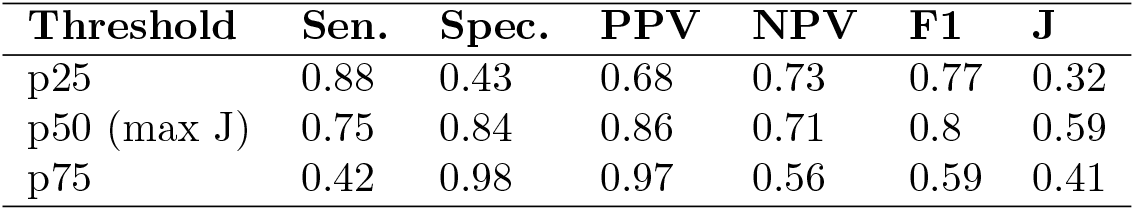
Evaluation with H12O’s dataset: performance metrics at different percentile thresholds. Percentiles 25%, 50% and 75% are selected, as well as the one that maximises Youden’s Index (J).

| Threshold | Sen. | Spec. | PPV | NPV | F1 | J |
| --- | --- | --- | --- | --- | --- | --- |
| p25 | 0.88 | 0.43 | 0.68 | 0.73 | 0.77 | 0.32 |
| p50 (max J) | 0.75 | 0.84 | 0.86 | 0.71 | 0.8 | 0.59 |
| p75 | 0.42 | 0.98 | 0.97 | 0.56 | 0.59 | 0.41 |

**Table 4:** Validation with LaFe’s dataset: performance metrics at different percentile thresholds. Percentiles 25%, 50% and 75% are selected, as well as the one that maximizes Youden’s Index (J).

| Threshold | Sen. | Spec. | PPV | NPV | F1 | J |
| --- | --- | --- | --- | --- | --- | --- |
| p25 | 0.85 | 0.37 | 0.6 | 0.68 | 0.71 | 0.22 |
| p50 | 0.66 | 0.68 | 0.7 | 0.64 | 0.68 | 0.35 |
| p75 | 0.36 | 0.87 | 0.76 | 0.55 | 0.49 | 0.23 |
| p60 (max J) | 0.57 | 0.79 | 0.75 | 0.62 | 0.65 | 0.36 |

The *sensitivity* (recall) of the “complications” class is approximately 75% which means that from the training population, the model has correctly detected around 75% of the patients presenting complications; this, with a PPV (Positive Predictive Value or *Precision*) of approximately 86% which is interpreted as the reliability of the model when it says that the observation leads to a complication, being correct 86% of the time.

Similarly, the model predicts a patient to remain in remission with a specificity of approximately 84% and an NPV (Negative Predictive Value) of 71%. This means that the model correctly identifies 84% of patients who are still in remission after 90 days. Conversely, predicting a patient as “in remission” is correct in 71% of cases (certainty).

A balance between both metrics (*sensitivity* and *specificity*) can be obtained with the Youden’s Index *J* described in equation 1. It could be interpreted as a benchmark to see if the model meets its 90-day forecast purpose and can vary between 0 and 1, complete separation by model inputs of the remitting and complicating populations results in *J=1* while complete overlap is *J=0* Fluss et al. (2005). The model achieves a *J* around 0.59.

### 3.4. External validation with LAFE dataset

An external validation is performed with a dataset unseen during model training and captured under different conditions in another hospital. The goal is to provide an estimate of the performance that could be expected when the model deals with data from new patients. This could answer if this model could be applied as it is to data coming from other healthcare centers or, on the contrary, it would be necessary to expand the dataset by combining data in a multicentric way.

Some features in the external dataset must be rescaled to match the units in the H12O dataset. Additionally, a subset of 23 features are present in both datasets, so the remaining 13 that cannot be found in LAFE are imputed in their entirety. Figure 5 presents the sensitivity-specificity curve obtained when the model (trained with data from H12O) is evaluated with data from La Fe, with an associated ROC-AUC of 0.7.

**Figure 5:**
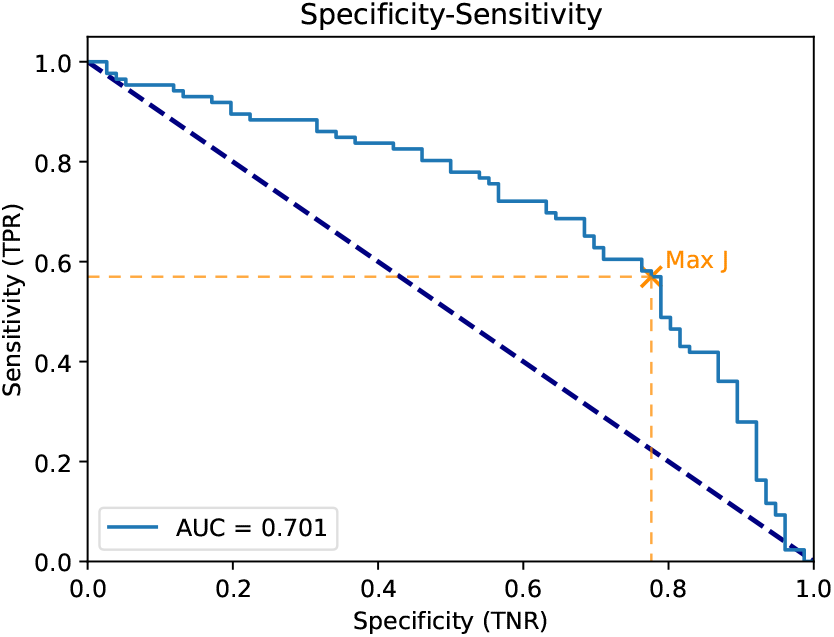
Sensitivity-specificity curve for the validation dataset. The operating point is superimposed for maximum Youden’s index (J) at 0.45 a-posteriori probability, percentile 60%.

### 3.5. Datasets comparison

In order to explain the loss of predictive power in the test data (collected under conditions other than those used for training), the similarity of the distributions of each variable is measured using the Jensen-Shannon divergence (JSD) Lin (1991). This metric equals 0 when the distributions are identical and tend towards 1 as divergence increases. Table 5 ranks the variables by decreasing JSD and relates it to the feature importance presented in Figure 3. It can be observed that JAK2, TP53 and Age are important variables that exhibit the highest distribution dissimilarities between train and test. Also, wbc, the second variable by importance, shows an intermediate divergence. These observations could explain part of the loss of predictive power (or lack of generalisation).

**Table 5:** Common variables between LAFE and H12O ranked by decreasing Jensen-Shannon Divergence.

|  | JSD | Importance rank |
| --- | --- | --- |
| JAK2_VAF | 0.29 | 5 |
| TP53_VAF | 0.27 | 4 |
| Age | 0.26 | 1 |
| TET2_VAF | 0.21 | 8 |
| KIT_VAF | 0.21 | 9 |
| wbc | 0.17 | 2 |
| CBL_VAF | 0.17 | 10 |
| Gender | 0.11 | 3 |
| minus7_7Q | 0.02 | 6 |

The distributions of the three most divergent variables and wbc are presented in figure 6.

**Figure 6:**
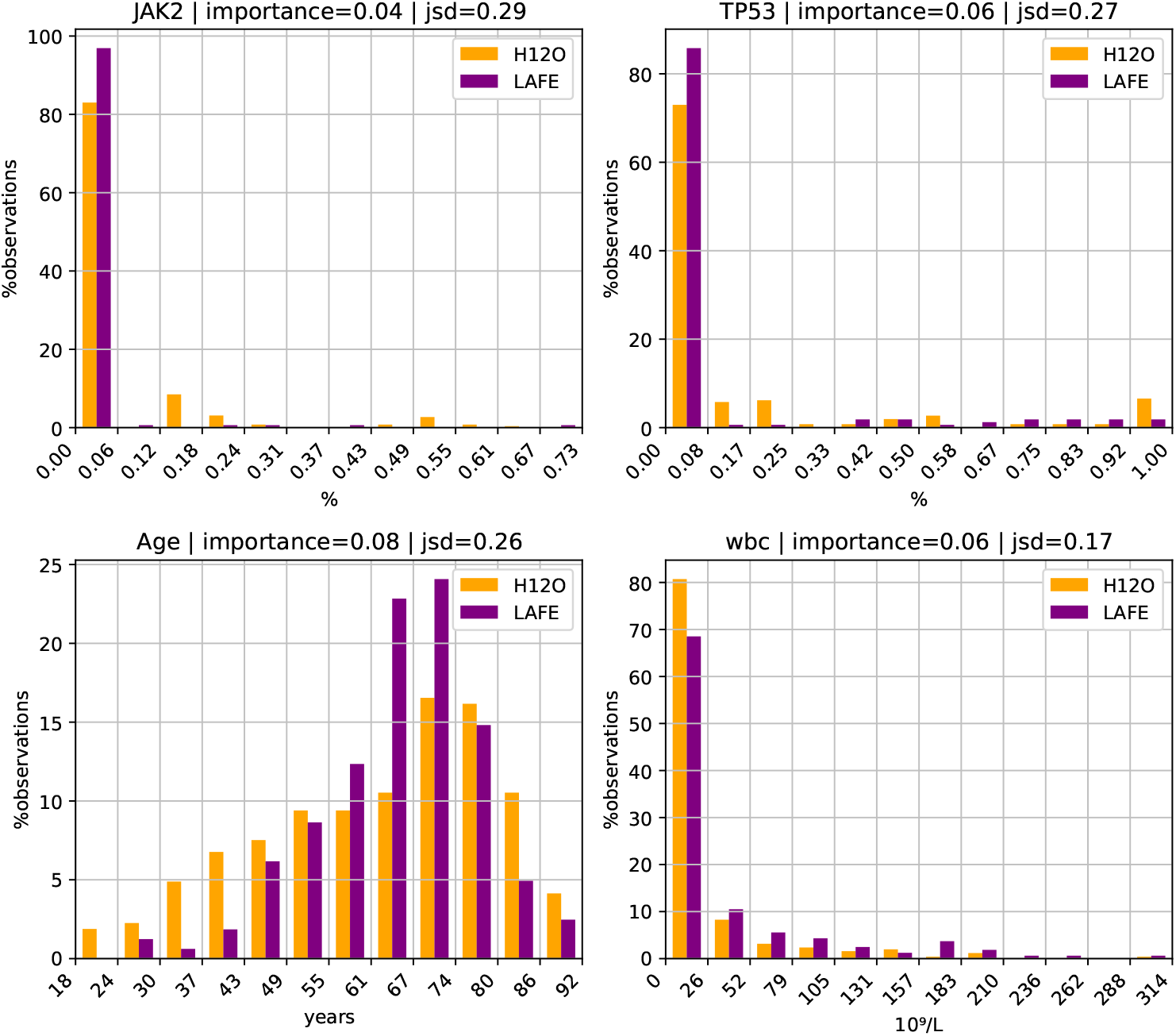
Comparison of the distributions of the first four most important variables. (orange) training cohort H12O. (purple) test population LAFE. (x-axis) variable value bins. (y-axis) for each bin, percentage of patients.

## 4. Discussion

In this work, we have developed a model for predicting 90-day mortality and response to treatment based on clinical and biological characteristics at diagnosis. This model has been validated in a set of observations, including fit and non-fit patients for intensive treatment. Unlike other models which only aim to predict early mortality Siddiqui et al. (2022), our approach has the advantage of taking into consideration mortality as well as treatment failure and recurrence, all accounting for possible complications. The resulting ROC-AUC (85% with original data, 70% with external validation data) indicates an overall satisfactory performance. A model designed with this approach in mind allows us to detect instances where a patient could be given an alternative treatment due to an estimated high risk of complications with the standard procedure.

This satisfactory predictive power could be due to the inclusion of biological, AML-related characteristics of the patient: leukocyte counts, cytogenetic alterations associated with an adverse prognosis and the mutational load from a large panel of genes with suspected implication in AML. Of the 27 VAF features present in the final model (Table 2), 9 are included in the ELN 2022 risk classification Döhner et al. (2022): CEBPA, EZH2, FLT3, KMT2A, RUNX1, SF3B1, SRSF2, TP53 and ZRSR2. The most important genetic feature in the model (TP53 VAF, as seen in Figure 3) is also present in the previously mentioned list. Most of these genes have been reported to have an adverse impact. It is worth noting that our model does not incorporate the presence of the mutated gene but the mutational load (in a continuous spectre) of the specific genes. This mutational load demonstrates its prognostic value, which leads us to consider if this data should be included in the prognostic classifications. The prognostic value of mutational load was already demonstrated in a previous work within the FLUGAZA trial with patients unfit for intensive treatment Ayala et al. (2021). Regarding cytogenetic abnormalities, all 5 present in the model (*t(8;21), inv(16)/t(16;16), -5/5q, -7* and *-17/17p*) are also present in the ELN 2022 classification.

Other models have included gene expression profiling (or transcriptomics) of 17 genes Huang et al. (2022) in a cohort of recurrence or refractory pediatric acute myeloid leukemia. Their results improve with patients with mutations in genes like FLT3-ITD, CEBPA or KMT2A rearrangements, all of which are considered inside our final model.

While the leave-one-out performance with the H12O data reaches a ROC-AUC of 0.85, the performance drops to 0.7 when validating with the external data of LAFE. We have found several factors that could explain this change in performance. Firstly, while the 36 features included in the final model are a subset of the extensive dataset of H12O, the LAFE’s dataset has only 23 common features directly usable by the model. The other 13 variables are imputed. Fortunately, the four most important features are present in both datasets. Another source of explanation to be considered concerns the differences in distribution between populations. In Section 3.5, slight differences in important variables *age, white-blood-cell count, TP53* and *gender* were observed. Furthermore, there is a difference with *a priori* probabilities: while 57.5% of the patients suffer complications at 90 days in H12O, this number drops to 53% in LAFE. With all this in mind, the drop in performance was to be expected, and poor generalisation power of the model should not be entirely blamed.

## 5. Conclusions

In this work, an XGBoost model trained with variables available at diagnosis has been proposed to predict the 90-day risk of complications after initiating the treatment for AML. Variant allele frequencies of several candidate genes, as well as cytogenetic abnormalities and other demographic variables have been gathered in two hospitals (H12O and LAFE).

To ensure the availability of a separate testing set, the model has been optimised and trained only with observations from H12O. Training and validating with data from this same institution with cross-validation achieved satisfactory results (0.85 ROC-AUC), with age, white blood cell counts and TP53 gene VAF being found as the most important predictors. Many of the genetic abnormalities included as features in the final model are classified as relevant in the 2022 European LeukemiaNet.

Regarding the external validation, the test results with data from LAFE yielded lower results (0.7 ROC-AUC), which can be due to several different causes, such as the observed differences in important variables distributions and the absence of some predictor variables in the test set. Despite this, the test metrics observed at the assigned operating point (0.75 PPV, 0.62 NPV) suggest that this model could be used as a therapeutical decision support tool. The reported model performance, especially with external validation data, would be expected to improve when given a dataset of better quality that caters to the expected features of the trained model.

## 6. Future work

Future work proposals are listed below:

- Compare the model’s performance varying the period of prediction (n-days). The more one wishes to anticipate the risk of long-term complications, the greater the uncertainty (loss of performance). In practice, a model with the focus set in a longer period after diagnosis, e.g. one year, could be as useful as the one presented in this work. This solution could come in the form of several classification models, each one trained with different n-days, or a single regression (survival) model which predicts the risk of complications for a patient at any given point in time.
- Identify relevant features with an alternative, more robust method that takes into consideration multivariate interactions. Knowing the subset of features that yields the best separation between classes could allow making more affordable predictions, improve model efficiency and expand our knowledge about AML.
- Improving the quality of the currently available datasets would allow us to incorporate more variables into the model. Furthermore, adding more patients into the study could strengthen model convergence. Gathering data from multiple institutions could help alleviate possible problems in model generalisation by accounting for potential issues such as patient populations of different characteristics being sampled or using different measurement tools.

## Data Availability

All data produced in the present study are available upon reasonable request to the authors.

## Acknowledgements

This work was partially supported by the Instituto de Salud Carlos III (ISCIII) through the projects PI19/00730, PI19/01518 and PI22/1088. It was co-funded by the European Union and by the Instituto de Investigación Hospital 12 de Octubre (imas12).

This work was also funded by Generalitat Valenciana through IVACE (Valencian Institute of Business Competitiveness, https://www.ivace.es/index.php/es/) and the European Union through FEDER funding under project IMDEEA/2022/23.

## Notes

### Competing Interest Statement

The authors have declared no competing interest.

### Author Declarations

The Institutional Ethics Committee for Clinical Research of Instituto de Investigación Sanitaria La Fe (IISLaFe, Valencia, Spain) approved this study with registration number 2019/0117. The Comité de Ética de la Investigación Clínica con Medicamentos (CEIm) of the Hospital 12 de Octubre (Madrid, Spain) approved this work with registration number 19/434.

